# Scleral Allograft-Reinforced Cyclodialysis Combined with Phacoemulsification or Performed as a Standalone Procedure: 24-Month Outcomes from the Multicenter CREST Study

**DOI:** 10.64898/2026.11.02.26367111

**Authors:** Robert N. Weinreb, Lorraine Provencher, Alex S. Huang, Arkadiy Yadgarov, Matthew Rouse, Brian Francis, Tsontcho Ianchulev, Robert Chang

## Abstract

**Introduction:** Acellular scleral allograft-reinforced cyclodialysis is a bio-interventional surgery (BIOS) to lower intraocular pressure (IOP). We report 2-year efficacy and safety outcomes when the procedure is performed in combination with phacoemulsification and as a standalone procedure in eyes with open-angle glaucoma.

**Methods:** Analysis of the multicenter CREST real-world evidence study of eyes undergoing allograft-reinforced cyclodialysis with AlloFlo™ Uveo (Iantrek, Inc., White Plains, NY; NCT05506423). Eyes attending the Month-24 visit were analyzed: 176 eyes undergoing phaco-combined surgery (Cohort 1) and 32 standalone eyes (Cohort 2). Values recorded after secondary procedures censored. Cohort 1 only was stratified by low, moderate, or high baseline IOP (≤ 15, 16–20, ≥ 21 mmHg).

**Results:** A total of 176 eyes in Cohort 1 and 32 in Cohort 2 were analyzed. In Cohort 1, IOP decreased 3.1 mmHg at Month 24 from a baseline of 18.0 (−12.5%; p < 0.001), and medications decreased 0.5 from 1.5 (p < 0.001); medication-free eyes rose from 22.9% to 46.4% (p < 0.001). IOP changes in the low, moderate, and high baseline tiers were +0.9 (+7.8%), −3.0 (−16.3%), and −9.2 (−36.1%) mmHg; medication changes were −0.8, −0.6, and +0.1. In Cohort 2, IOP decreased 6.4 mmHg from 20.0 (−27.1%; p < 0.001), and medications decreased 1.2 from 2.8 (p = 0.004); medication-free eyes rose from 4.3% to 30.4% (p = 0.031). Secondary IOP-lowering procedures were performed in 5.1% of Cohort 1 eyes and 28.1% of Cohort 2. Kaplan–Meier freedom from reoperation was 98.9% and 84.4%. No eye developed clinically significant hypotony or choroidal effusion, and there were no allograft-related vision-threatening adverse events.

**Conclusion:** Allograft-reinforced cyclodialysis effectively lowered IOP and medication use through 24 months.

**Trial Registration:** ClinicalTrials.gov NCT05506423.

**Plain Language Summary:** Glaucoma damages the optic nerve, usually because pressure inside the eye is too high. A newer operation opens a small channel to a natural drainage route inside the eye and holds it open with thin pieces of biocompatible donor tissue. It spares the surface of the eye, so other operations remain possible later. We asked how well the procedure works after two years. We studied 208 eyes at 17 centers that returned for their two-year visit: 176 done with cataract surgery and 32 done as a standalone procedure. We measured eye pressure and drop use and checked for long-term safety. With cataract surgery, pressure decreased from about 18 to 15 mmHg (millimeters of mercury, the standard measurement for eye pressure) and drop use by about half a drop; eyes starting at the highest pressures improved most, falling about 9 mmHg. Done alone, pressure decreased from about 20 to 14 mmHg and drop use by about one drop. The share of eyes needing no drops at all roughly doubled with cataract surgery, from about a quarter to about a half, and rose from almost none to about a third when done alone. No eye developed dangerously low pressure or fluid behind the retina. About one-in-four eyes treated alone needed a further pressure-lowering procedure within two years, against about one-in-twenty treated with cataract surgery. The procedure meaningfully lowered pressure and drop use over two years without the complications that ended earlier versions of this promising surgery.

**Key Summary Points:** *Why carry out this study?:*

- Eyes needing cataract surgery with better pressure control, and eyes needing pressure control alone, are managed very differently, yet 2-year data on allograft-reinforced cyclodialysis in both settings have not been reported together.
- We asked: how much do pressure and medication burden fall by 24 months in each setting, how often further surgery is needed, and how safe is it?

*What was learned from the study?:*

- When performed at the same time as cataract surgery, pressure decreased 3.1 mmHg and medications by 0.5, and the reductions rose steeply with baseline pressure. The proportion of eyes on no glaucoma medication doubled, from 22.9% to 46.4%.
- When performed as a standalone procedure, pressure decreased 6.4 mmHg and medications by 1.2, and medication-free eyes rose from 4.3% to 30.4%, reflecting promising outcomes in refractory eyes.
- No eye developed clinical hypotony, choroidal effusion, or corneal decompensation, and endothelial cell loss was confined to the phacoemulsification-combined cohort. Freedom from reoperation at 24 months was 98.9% with cataract surgery and 84.4% standalone.

## Introduction

Cataract and glaucoma frequently coexist. Lens extraction alone lowers intraocular pressure (IOP) by approximately 2–4 mmHg in glaucomatous eyes, with greater reductions associated with higher preoperative IOP [1–5]. Phacoemulsification combined with minimally invasive glaucoma surgery (MIGS) is routinely performed when additional IOP control or a reduction in medication burden is warranted. The incremental efficacy of the most widely used trabecular and canalicular devices over phacoemulsification alone is modest: pivotal randomized trials demonstrated between-group differences of 1.6–2.3 mmHg in unmedicated diurnal IOP reduction at 24 months [6, 7]. Canal interventions are mostly proximal to the collector channels and episcleral veins, and their pressure-lowering efficacy is therefore constrained by distal outflow resistance and episcleral venous pressure.

The uveoscleral pathway permits aqueous humor to traverse the ciliary interstitium into the supraciliary and suprachoroidal spaces, without the lower pressure limit imposed by episcleral venous pressure. Surgical cyclodialysis accesses this pathway, but its outcomes have historically been unpredictable. Cyclodialysis clefts are susceptible to spontaneous postoperative closure, which can cause IOP spikes and diminish the pressure-lowering effect. Cleft reinforcement is therefore indicated to maintain patency and provide sustained IOP reduction.

Scleral allograft-reinforced cyclodialysis entails creation of a controlled ab-interno cyclodialysis cleft and placement of shaped segments of minimally modified homologous, non-resorbable human scleral allograft within the supraciliary space to maintain cleft patency [8]. Published studies have reported first-in-human results, 12-month multicenter outcomes, safety outcomes in a large clinical series, single-center 24-month outcomes, and 12-month outcomes following standalone surgery [8–12]. However, multicenter safety and efficacy outcomes at 24 months have not yet been reported.

We report 24-month outcomes from two parallel cohorts of the CREST registry: phacoemulsification combined with allograft-reinforced cyclodialysis in phakic eyes (Cohort 1), and standalone allograft-reinforced cyclodialysis (Cohort 2). The cohorts answer different clinical questions and differ in lens status, baseline IOP, and referral pattern, so they are characterized in parallel and not compared with each other. Because IOP dependence on baseline pressure is a recurrent theme in MIGS reporting [5, 13], we examined it in the combined Cohort 1 as an exploratory analysis [14]. Enrollment numbers in the standalone cohort did not support subgroup stratification in the standalone Cohort 2.

## Methods

### Study Design and Participants

CREST (NCT05506423) is a prospective, multicenter, observational real-world evidence study that enrolls recipients of allograft-reinforced cyclodialysis using the AlloFlo Uveo™ system (Iantrek, Inc., White Plains, NY) at 17 sites in the United States and Central America. The study includes broad real-world inclusion criteria (diagnosis of open-angle glaucoma and bio-reinforced cyclodialysis was attempted) with no exclusion criteria. Per-protocol, eyes are enrolled into one of two cohorts: Cohort 1 comprises eyes that underwent the study procedure concurrent with phacoemulsification in a single surgical session; Cohort 2 comprises eyes that underwent a standalone study procedure. As a real-world study, there is no medication washout, and treating surgeons have postoperative discretion to medicate eyes according to need and clinical best practice. Likewise, there are no postoperative medication or treatment policies in the CREST protocol.

Eyes in the present analysis were required to have had at least 24 months of post-surgical follow-up to allow for adequate long-term analysis of efficacy and safety. The present analysis reports outcomes as of the most recent data cutoff (31 January 2026). The CREST study is conducted in accordance with the Declaration of Helsinki, under oversight of Advarra IRB (approval MOD02608603). All participants gave written informed consent.

### Surgical Procedure

The ab-interno bio-interventional surgical (BIOS) technique has been previously described [8, 9]. Briefly, under topical or intracameral anesthesia, a cyclodialysis cleft is created in the supraciliary space and then acellular scleral allograft bio-scaffolds are placed in the cleft. The allograft spacer is composed of microtrephined scleral donor tissue (5 mm × 500 µm each), rather than a rigid polymer, and the procedure preserves the conjunctiva and does not result in a filtering bleb. Allograft tissue is processed in a minimally modified standard and comes preloaded in a handheld delivery cannula. The number of segments and quadrant were at the treating surgeon’s discretion. Postoperative medications and the decision to withhold or continue glaucoma medications followed each surgeon’s best practice for the individual patient and eye.

### Measurements

IOP was measured by Goldmann applanation tonometry. Baseline IOP was the medicated IOP at the preoperative visit on the patient’s current regimen. Medications were counted by active ingredient, with fixed combinations counted as two and oral carbonic anhydrase inhibitors counted as one. Best-corrected visual acuity (BCVA) was converted to logMAR. When performed per clinician judgement, endothelial cell density (ECD) was captured and measured by specular microscopy. No reading center was employed, and measurements were verified for quality in the study’s data management process. Allograft position was assessed by gonioscopy.

### Outcomes

Effectiveness outcomes were: mean changes in medicated IOP and number of medications from baseline to Month 24 in each cohort, the proportions of eyes with ≥ 20% IOP reduction and with IOP ≤ 18 and ≤ 15 mmHg, and composite responders achieving the same IOP thresholds without increase in medications from baseline or secondary IOP-lowering procedures. Additional outcomes assessed were rates of secondary IOP-lowering procedures, change in visual acuity, and change in ECD. Time to reoperation was estimated by the Kaplan–Meier method with complementary log-log confidence intervals. Eyes without an event were censored at their last recorded study visit.

Safety outcomes were prospectively defined adverse events: clinical hypotony, choroidal effusion or folds, hyphema > 2 mm, loss of ≥ 2 lines of visual acuity at ≥ 90 days, IOP elevation ≥ 10 mmHg above baseline > 30 days after surgery, macular edema, confirmed visual field mean deviation worsening ≥ 2.5 dB, retinal complications, uveitis, and unplanned surgical reintervention, in addition to all other ocular adverse events.

### Handling of Secondary Procedures

Two definitions are distinguished and reported separately throughout. A secondary IOP-lowering procedure is any procedure performed to provide sustained IOP reduction and is the event that governs censoring for efficacy assessments. Anterior chamber paracentesis and bleb needling or revision are not counted as censoring procedures; all other IOP-lowering procedures result in censoring at the time they are performed. Reoperation counts incisional surgeries only, excluding laser procedures, and is the endpoint of the Kaplan–Meier analysis.

The primary analysis used a while-on-treatment strategy, excluding values recorded after a secondary procedure, so efficacy estimates describe eyes managed with the index procedure and medication without confounding from subsequent procedures. Composite responder analyses were calculated on all eyes attending Month 24, with eyes undergoing a secondary procedure counted as non-responders. A sensitivity analysis was performed in which eyes with a secondary IOP-lowering procedure were assigned their baseline IOP and medication count and classed as non-responders.

### Statistical Analyses

Continuous variables, including number of medications, are summarized as mean ± standard deviation (SD), and IOP is reported as mean with 95% confidence interval (CI). Within-cohort changes from baseline were tested with the paired t-test, the change in the proportion of medication-free eyes with McNemar’s exact test, and confidence intervals for mean changes and for mean percentage changes are t-based. Both eyes of some patients were included, and all data were analyzed at the eye level without adjustment for inter-eye correlation [15].

Because baseline medicated IOP in Cohort 1 ranged from 9 to 32 mmHg, eyes were stratified by baseline medicated IOP and analyzed in three tiers: low (≤ 15 mmHg), moderate (16–20 mmHg) and high (≥ 21 mmHg). A single pooled mean would otherwise blend eyes operated to lower pressure with eyes already at target operated to reduce medication burden. Missing data were handled by complete-case analysis without imputation. Analyses were performed in R version 4.5.3 (R Foundation for Statistical Computing).

## Results

### Baseline Characteristics

A total of 176 eyes in Cohort 1 and 32 in Cohort 2 met the eligibility criteria and were included for analysis. Before Month 24, 9 eyes in each cohort underwent a secondary IOP-lowering procedure; these eyes were excluded from the primary efficacy analyses, leaving N=167 and N=23 eyes in Cohort 1 and 2, respectively for efficacy analysis. These eyes were retained in the sensitivity, time-to-event, and safety analyses.

Demographics and baseline characteristics are shown in Table 1. Cohort 1 eyes (all phakic) had a mean age of 71.2 ± 7.5 years, 51.1% were female and 48.9% male, and mean baseline IOP was 17.9 mmHg (17.2, 18.6) on 1.4 ± 1.3 medications. Most eyes with visual field data (54.1%) had mild disease. Baseline IOP tiers comprised 35.8% of eyes in the low tier (≤ 15 mmHg), 40.3% moderate (16–20 mmHg), and 23.9% high (≥ 21 mmHg). Low-tier eyes carried the heaviest medication burden (1.9 ± 1.5 medications), consistent with pressure controlled by medication.

**Table 1.** Demographics and baseline disease characteristics.

| Demographic | Cohort 1<br>(N = 176) |  |  |  | Cohort 2<br>(N = 32) |
|---|---|---|---|---|---|
|  | Overall<br>(N = 176) | Low,<br>IOP ≤ 15 mmHg<br>(N = 63) | Moderate,<br>IOP 16–20 mmHg<br>(N = 71) | High,<br>IOP ≥ 21 mmHg<br>(N = 42) |  |
| Age, mean years ± SD | 71.2 ± 7.5 | 70.4 ± 8.7 | 71.7 ± 6.9 | 71.8 ± 6.7 | 73.8 ± 8.5 |
| Sex, n (%) |  |  |  |  |  |
| Female | 90 (51.1%) | 39 (61.9%) | 33 (46.5%) | 18 (42.9%) | 19 (59.4%) |
| Male | 86 (48.9%) | 24 (38.1%) | 38 (53.5%) | 24 (57.1%) | 13 (40.6%) |
| Race and ethnicity, n (%) |  |  |  |  |  |
| White | 117 (66.5%) | 41 (65.1%) | 47 (66.2%) | 29 (69.0%) | 26 (81.3%) |
| Black | 56 (31.8%) | 22 (34.9%) | 21 (29.6%) | 13 (31.0%) | 5 (15.6%) |
| Asian | 3 (1.7%) | 0 | 3 (4.2%) | 0 | 0 |
| Hispanic or Latino | 35 (19.9%) | 5 (7.9%) | 16 (22.5%) | 14 (33.3%) | 9 (28.1%) |
| **Baseline disease characteristic** |  |  |  |  |  |
| Phakic lens status | 176 (100%) | 63 (100%) | 71 (100%) | 42 (100%) | 1 (3.1%) |
| Severity, mean VF MD, dB ± SD□ | −7.2 ± 7.4 | −7.7 ± 8.4 | −6.7 ± 6.1 | −7.0 ± 7.9 | −12.8 ± 7.3 |
| Mild, > –6 dB, n (%) | 59 (54.1%) | 25 (55.6%) | 21 (50.0%) | 13 (59.1%) | 5 (21.7%) |
| Moderate, –6 to –12 dB, n (%) | 20 (18.3%) | 7 (15.6%) | 10 (23.8%) | 3 (13.6%) | 6 (26.1%) |
| Severe, < –12 dB, n (%) | 30 (27.5%) | 13 (28.9%) | 11 (26.2%) | 6 (27.3%) | 12 (52.2%) |
| Prior procedures, n (%) |  |  |  |  |  |
| SLT | 35 (19.9%) | 18 (28.6%) | 12 (16.9%) | 5 (11.9%) | 17 (53.1%) |
| MIGS | 0 | 0 | 0 | 0 | 10 (31.3%) |
| Incisional surgery | 2 (1.1%) | 2 (3.2%) | 0 | 0 | 7 (21.9%) |
| Bleb-forming surgery | 1 (0.6%) | 1 (1.6%) | 0 | 0 | 7 (21.9%) |
| Baseline IOP, mmHg (95% CI) | 17.9 (17.2, 18.6) | 13.3 (12.9, 13.7) | 17.9 (17.6, 18.2) | 24.9 (24.0, 25.9) | 20.8 (18.9, 22.8) |
| Medications, mean ± SD | 1.4 ± 1.3 | 1.9 ± 1.5 | 1.3 ± 1.1 | 1.0 ± 1.2 | 2.9 ± 1.2 |
| Medication free, n (%) | 42 (23.9%) | 7 (11.1%) | 16 (22.5%) | 19 (45.2%) | 1 (3.1%) |
| BCVA, logMAR ± SD<sup>b</sup> | 0.34 ± 0.44 | 0.27 ± 0.35 | 0.30 ± 0.39 | 0.50 ± 0.59 | 0.41 ± 0.64 |

BCVA = best-corrected visual acuity; CI = confidence interval; IOP = intraocular pressure; MD = mean deviation; MIGS = minimally invasive glaucoma surgery; SD = standard deviation; SLT = selective laser trabeculoplasty; VF = visual field.
a. Disease severity based on visual field mean deviation in dB: mild > –6 dB; moderate –6 to –12 dB; severe < –12 dB. Baseline visual field MD was available for N = 109 in Cohort 1 (by tier N = 45 / 42 / 22) and N = 23 in Cohort 2.
b. Baseline BCVA available for N = 169 in Cohort 1 (by tier N = 60 / 69 / 40) and N = 25 in Cohort 2. Nine eyes had acuities recorded off the chart; these were assigned conventional logMAR equivalents: counting fingers 1.9, hand movements 2.3, no light perception 3.0.

Cohort 2 eyes had a mean age of 73.8 ± 8.5 years, 59.4% were female and 40.6% male, 96.9% were pseudophakic, and mean baseline IOP was 20.8 mmHg (18.9, 22.8) on 2.9 ± 1.2 medications. The baseline IOP distribution in Cohort 2 was 12.5% low, 31.3% moderate, and 56.3% high; these proportions are descriptive only, and the stratified analysis was confined to Cohort 1, which was the only cohort large enough to support tier-level estimates.

### Efficacy in Cohort 1: Combined Phacoemulsification and Bio-reinforced Cyclodialysis

IOP and medication counts over time are shown in Fig. 1. In the 167 eyes of the per-protocol primary efficacy analysis, mean IOP decreased from 18.0 mmHg (17.2, 18.7) to 14.9 mmHg (14.3, 15.4), an absolute change of −3.1 mmHg (−3.9, −2.2) and a relative change of −12.5% (−16.7%, −8.3%; p < 0.001; Table 2). Mean medication count among 166 eyes with paired data decreased from 1.5 ± 1.3 to 1.0 ± 1.2 (−0.5 ± 1.1; p < 0.001). The proportion of medication-free eyes rose from 22.9% to 46.4% (+23.5 percentage points; p < 0.001): 52 eyes became medication-free, while 13 that had been medication-free at baseline were taking medication at Month 24.

**Fig. 1.**
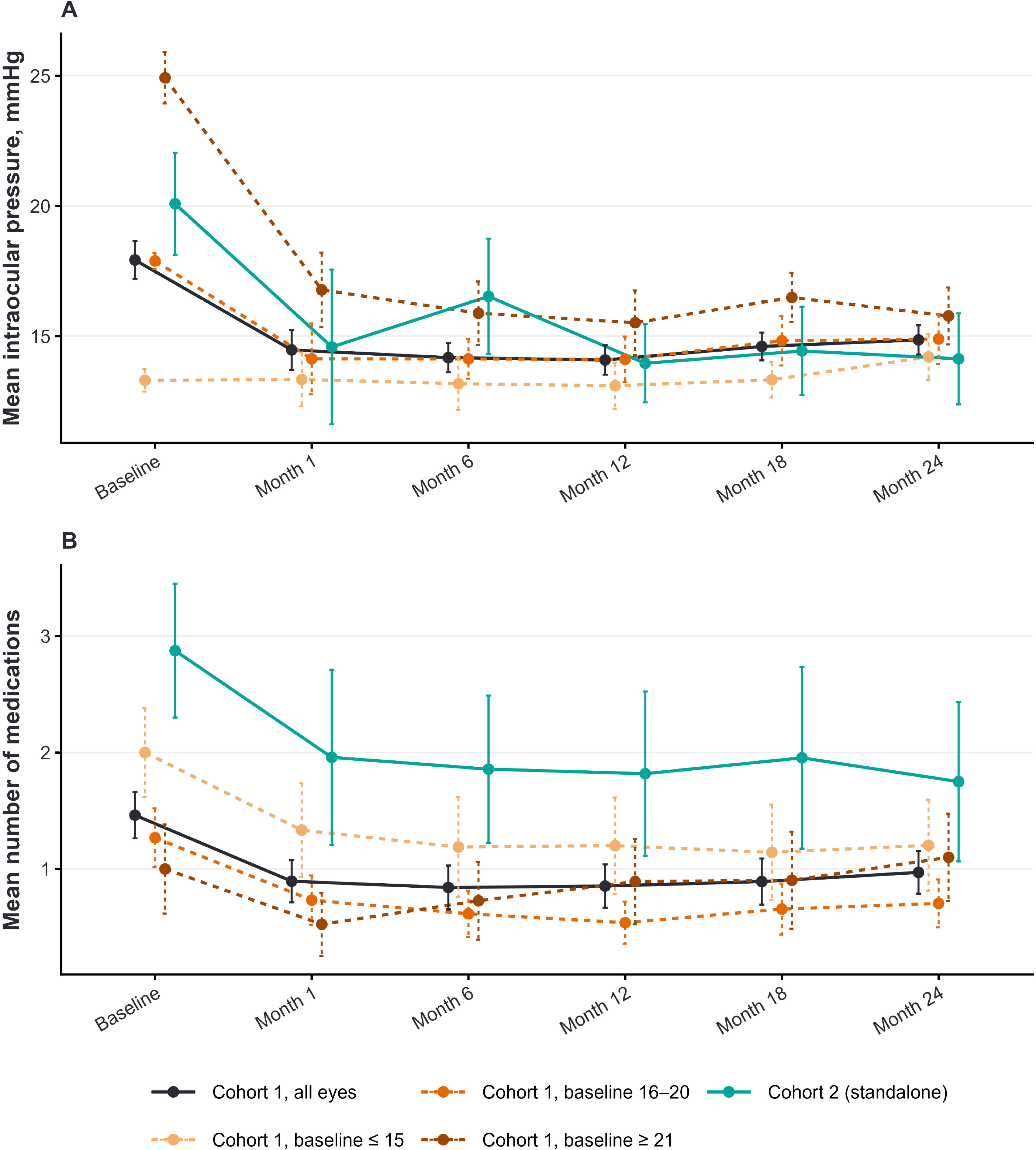
Mean intraocular pressure (IOP) (A) and medication count (B) with 95% confidence intervals (CIs) at each scheduled visit from baseline to Month 24, by cohort and, for Cohort 1, by baseline tier.

**Table 2.** Intraocular pressure and medication outcomes at Month 24.

| Efficacy Analysis, units | Cohort 1<br>(N = 167) |  |  |  | Cohort 2<br>(N = 23) |
|---|---|---|---|---|---|
|  | Overall<br>(N = 167) | Low,<br>IOP ≤ 15 mmHg<br>(N = 59) | Moderate,<br>IOP 16–20 mmHg<br>(N = 68) | High,<br>IOP ≥ 21 mmHg<br>(N = 40) |  |
| **IOP primary analysis: post-secondary-procedure values censored** |  |  |  |  |  |
| M24 IOP, mmHg (95% CI) | 14.9 (14.3, 15.4) | 14.3 (13.4, 15.2) | 14.9 (13.9, 15.9) | 15.8 (14.7, 16.9) | 13.7 (12.1, 15.2) |
| IOP change, mmHg (95% CI) | −3.1 (−3.9, −2.2) | +0.9 (+0.1, +1.8) | −3.0 (−4.0, −2.0) | −9.2 (−10.5, −7.8) | −6.4 (−9.0, −3.8) |
| Relative change, % (95% CI) | −12.5% (−16.7%, −8.3%) | +7.8% (+1.4%, +14.3%) | −16.3% (−22.0%, −10.6%) | −36.1% (−40.7%, −31.5%) | −27.1% (−40.4%, −13.7%) |
| p-value | p < 0.001 | p = 0.028 | p < 0.001 | p < 0.001 | p < 0.001 |
| **Medication primary analysis: post-secondary-procedure values censored** |  |  |  |  |  |
| M24 medications, n ± SD | 1.0 ± 1.2 | 1.2 ± 1.5 | 0.7 ± 0.9 | 1.1 ± 1.2 | 1.6 ± 1.5 |
| Medication change, n ± SD | −0.5 ± 1.1 | −0.8 ± 0.9 | −0.6 ± 1.1 | +0.1 ± 1.2 | −1.2 ± 1.7 |
| p-value | p < 0.001 | p < 0.001 | p < 0.001 | p = 0.606 | p = 0.004 |
| Medication free at M24, n (%) | 77 (46.4%) | 28 (48.3%) | 35 (51.5%) | 14 (35.0%) | 7 (30.4%) |
| Change in med-free from BL<sup>a</sup> | +23.5% | +37.9% | +30.9% | −10.0% | +26.1% |
| p-value | p < 0.001 | p < 0.001 | p < 0.001 | p = 0.388 | p = 0.031 |
| **Responder analyses at Month 24: IOP thresholds use the censored sets; composites use all eyes attending Month 24** |  |  |  |  |  |
| IOP ≤ 18 mmHg, n (%) | 144 (86.2%) | 54 (91.5%) | 58 (85.3%) | 32 (80.0%) | 21 (91.3%) |
| Composite responder | 127 (72.2%) | 51 (81.0%) | 52 (73.2%) | 24 (57.1%) | 18 (56.3%) |
| IOP ≤ 15 mmHg, n (%) | 94 (56.3%) | 35 (59.3%) | 41 (60.3%) | 18 (45.0%) | 17 (73.9%) |
| Composite responder | 80 (45.5%) | 33 (52.4%) | 35 (49.3%) | 12 (28.6%) | 14 (43.8%) |
| ≥ 20% IOP reduction, n (%) | 75 (44.9%) | 6 (10.2%) | 34 (50.0%) | 35 (87.5%) | 16 (69.6%) |
| Composite responder | 61 (34.7%) | 5 (7.9%) | 29 (40.8%) | 27 (64.3%) | 13 (40.6%) |
| ≥ 30% IOP reduction, n (%) | 49 (29.3%) | 3 (5.1%) | 21 (30.9%) | 25 (62.5%) | 13 (56.5%) |
| Composite responder | 36 (20.5%) | 2 (3.2%) | 17 (23.9%) | 17 (40.5%) | 10 (31.3%) |
| **Sensitivity analysis: secondary procedures counted as failures, baseline IOP and medication counts carried forward** |  |  |  |  |  |
|  | **N = 176** | **N = 63** | **N = 71** | **N = 42** | **N = 32** |
| M24 IOP, mmHg (95% CI) | 15.0 (14.5, 15.6) | 14.2 (13.4, 15.0) | 15.0 (14.1, 16.0) | 16.2 (15.0, 17.4) | 16.2 (14.0, 18.4) |
| IOP change, mmHg (95% CI) | −2.9 (−3.7, −2.1) | +0.9 (+0.1, +1.7) | −2.9 (−3.8, −1.9) | −8.7 (−10.1, −7.3) | −4.6 (−6.7, −2.5) |
| Relative change, % (95% CI) | −11.9% (−15.9%, −7.8%) | +7.3% (+1.3%, +13.4%) | −15.6% (−21.1%, −10.1%) | −34.4% (−39.3%, −29.4%) | −19.5% (−29.8%, −9.1%) |
| p-value | p < 0.001 | p = 0.029 | p < 0.001 | p < 0.001 | p < 0.001 |
| M24 medications, n ± SD | 1.0 ± 1.2 | 1.2 ± 1.5 | 0.7 ± 0.9 | 1.1 ± 1.2 | 2.0 ± 1.5 |
| Medication change, n ± SD | −0.5 ± 1.1 | −0.8 ± 0.9 | −0.6 ± 1.1 | +0.1 ± 1.2 | −0.8 ± 1.6 |
| p-value | p < 0.001 | p < 0.001 | p < 0.001 | p = 0.605 | p = 0.005 |

BL = baseline; CI = confidence interval; IOP = intraocular pressure; M = month; SD = standard deviation.
Notes: Composite responders met the IOP threshold with no increase in medication count from baseline and no secondary IOP-lowering procedure. In the sensitivity analysis, eyes with a secondary IOP-lowering procedure were assigned their baseline IOP and medication count and classed as non-responders.
a. Analyzed among eyes with paired baseline and Month 24 counts (N = 166 / 58 / 68 / 40 / 23). Change from baseline is percentage points, tested with McNemar’s exact test.

Responder analyses showed that 144 eyes (86.2%) had IOP ≤ 18 mmHg and 94 (56.3%) ≤ 15 mmHg; 75 (44.9%) achieved ≥ 20% IOP reduction and 49 (29.3%) ≥ 30% (Fig. 3). On the composite definition, which additionally requires no increase in medication and no secondary procedure and is calculated on all 176 eyes, the corresponding proportions were 72.2%, 45.5%, 34.7% and 20.5%.

Kaplan–Meier freedom from reoperation at 24 months was 98.9% (95.5%, 99.7%) in Cohort 1 and 84.4% (66.5%, 93.2%) in Cohort 2 (Fig. 2; p < 0.001). Counting the 9 eyes with a secondary procedure as failures, the mean IOP change was −2.9 mmHg (−3.7, −2.1), 75 of 176 eyes (42.6%) achieved ≥ 20% reduction, and 151 (85.8%) had IOP ≤ 18 mmHg.

**Fig. 2.**
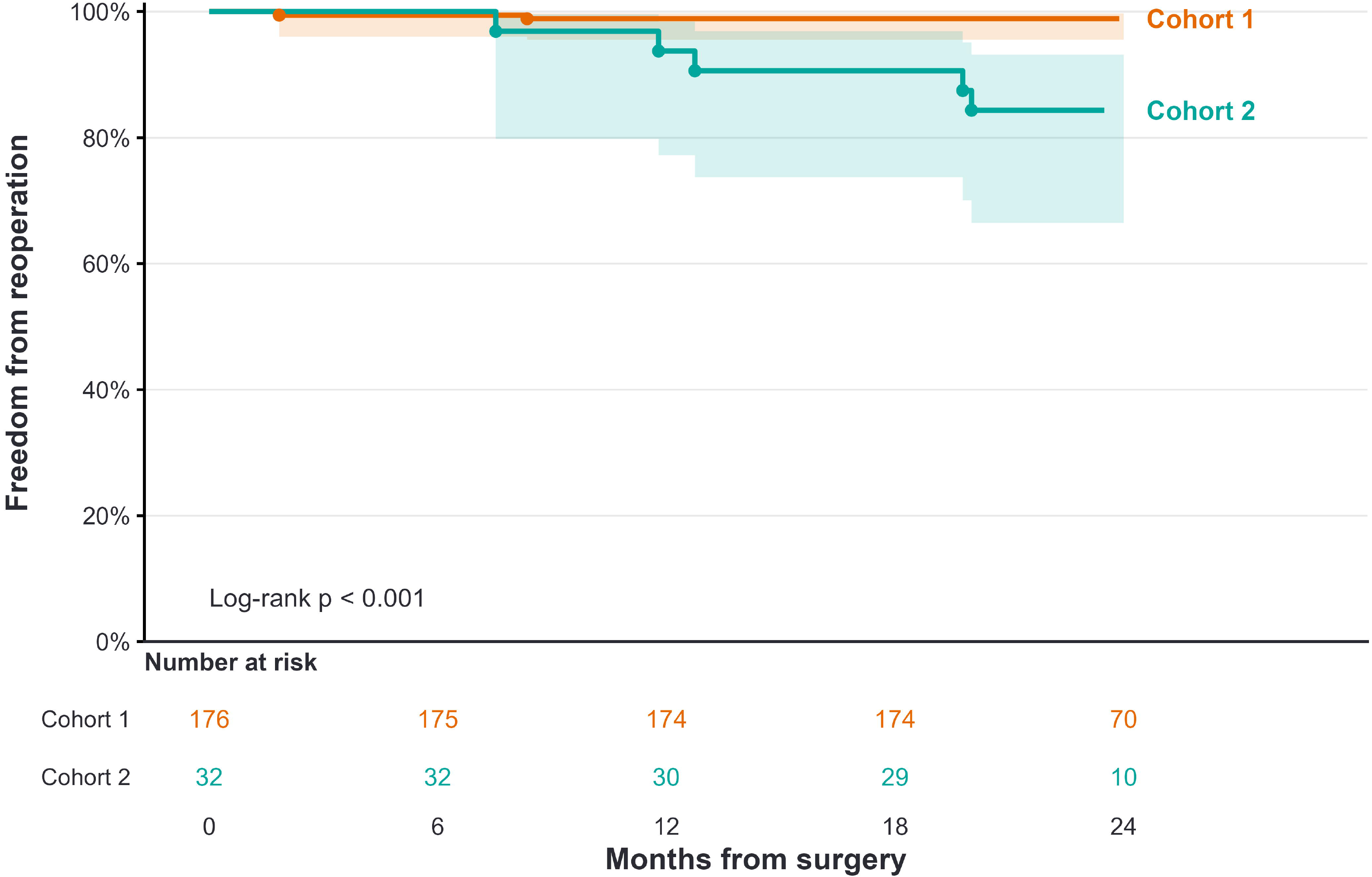
Kaplan–Meier freedom from reoperation for Cohort 1 and Cohort 2. Reoperation counts incisional IOP-lowering procedures only; laser trabeculoplasty, cyclophotocoagulation, anterior chamber paracentesis and bleb revision or needling are not counted. Eyes without an event are censored at their last recorded visit. Shaded bands are 95% confidence intervals (complementary log-log). Circles mark individual reoperations. Numbers at risk are shown beneath the time axis.

### Cohort 1 Analysis by Baseline IOP

IOP and medication changes by tier are shown together in Table 2. In the low tier (N = 59), IOP increased by 0.9 mmHg (+0.1, +1.8; p = 0.028) while medications decreased by 0.8 ± 0.9 (p < 0.001); 65.5% of eyes reduced medications, and 91.5% had IOP ≤ 18 mmHg at Month 24. In the moderate tier (N = 68), IOP decreased by 3.0 mmHg (−4.0, −2.0; −16.3%; p < 0.001) and medications decreased 0.6 ± 1.1 (p < 0.001). In the high tier (N = 40), IOP decreased by 9.2 mmHg (−10.5, −7.8; −36.1%; p < 0.001), with no change in medications (+0.1 ± 1.2; p = 0.606) and no significant change in the medication-free proportion (45.0% to 35.0%; p = 0.388). In the high-IOP tier, 87.5% achieved ≥ 20% reduction, 62.5% ≥ 30% reduction, and 80.0% had IOP reduced to ≤ 18 mmHg. In a sensitivity analysis in the high-IOP tier that counted eyes with a secondary procedure as failures, these proportions were 83.3%, 59.5% and 76.2%. Month 24 IOP exceeded 18 mmHg in 5 low-tier eyes (8.5%), all of which had a baseline IOP of 15 mmHg or less, in 10 moderate-tier eyes (14.7%), and in 8 high-tier eyes (20.0%). IOP increased by at least 5 mmHg in 6 low-tier eyes, all of which simultaneously reduced medications.

### Cohort 2: Standalone Allograft-Reinforced Cyclodialysis

In the 23 eyes of the primary analysis, mean IOP decreased from 20.0 mmHg (18.0, 22.1) at baseline to 13.7 mmHg (12.1, 15.2) at Month 24, a change of −6.4 mmHg (−9.0, −3.8; p < 0.001), a relative reduction of 27.1% (−40.4%, −13.7%; Fig. 1). Medications decreased from 2.8 ± 1.3 at baseline to 1.6 ± 1.5 at Month 24, a reduction of −1.2 ± 1.7 medications (p = 0.004); 52.2% of eyes were using fewer medications at Month 24 than they were at baseline, and medication-free eyes rose from 4.3% to 30.4% (p = 0.031), with 6 eyes becoming medication-free and none losing that status. A total of 16 eyes (69.6%) achieved a ≥ 20% reduction, 13 (56.5%) a ≥ 30% reduction, and 21 (91.3%) had IOP ≤ 18 mmHg and 17 (73.9%) had IOP ≤ 15 mmHg (Fig. 3). On the composite definition applied to all 32 eyes, the corresponding proportions were 40.6%, 31.3%, 56.3% and 43.8%. Kaplan–Meier freedom from reoperation at 24 months was 84.4% (66.5%, 93.2%; Fig. 2).

**Fig. 3.**
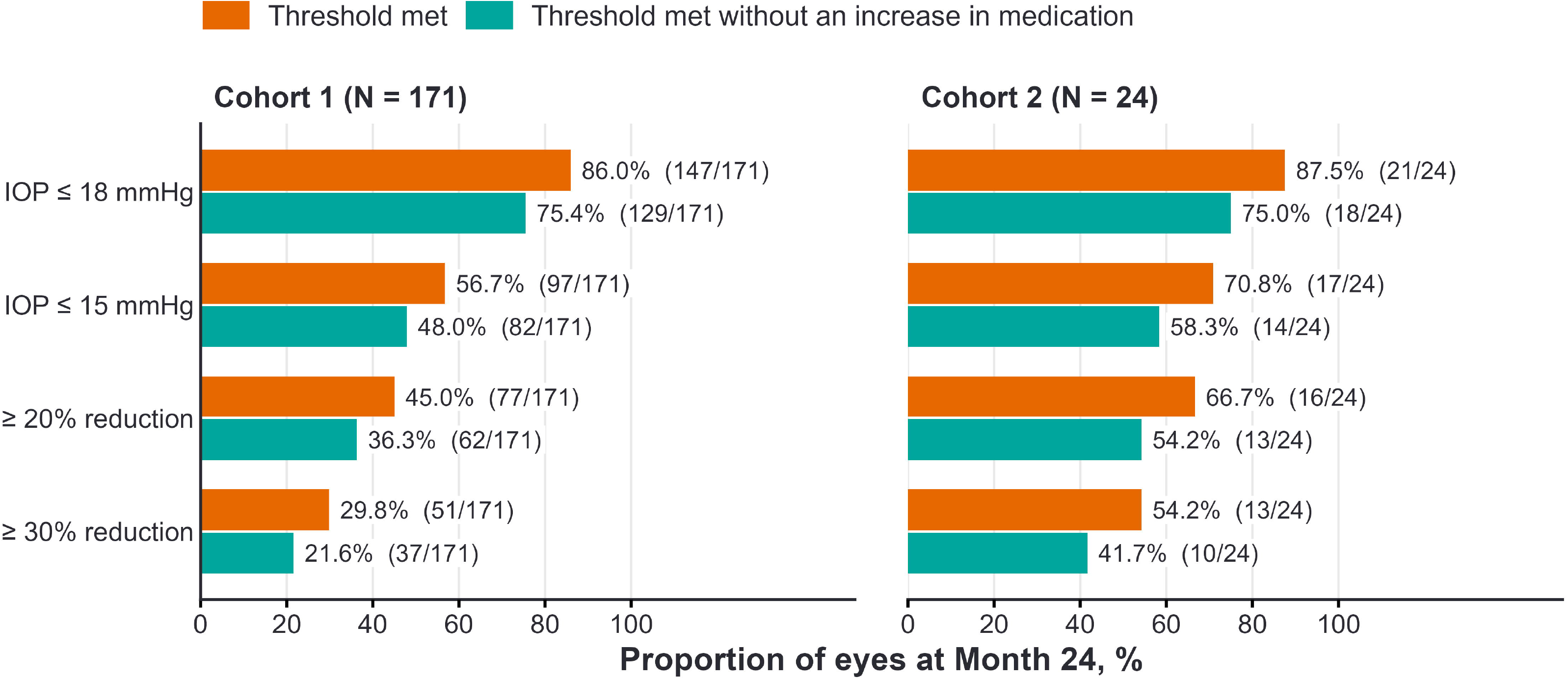
Proportion of eyes meeting each intraocular pressure threshold at Month 24, by cohort. Orange bars (threshold met) are calculated among eyes whose Month 24 pressure preceded any further IOP-lowering procedure: Cohort 1, N = 167; Cohort 2, N = 23. Teal bars (composite responder) require the same threshold to be met with no increase in glaucoma medication from baseline and no secondary IOP-lowering procedure, and are calculated among all eyes attending Month 24, with eyes undergoing a secondary procedure counted as non-responders: Cohort 1, N = 176; Cohort 2, N = 32.

In the sensitivity analysis, 9 eyes with a secondary procedure were counted as failures and their baseline IOP and medication counts carried forward. The Month 24 IOP change was −4.6 mmHg (−6.7, −2.5; −19.5%; p < 0.001) with 50.0% achieving a ≥ 20% reduction.

### Safety

Adverse events (AEs) are summarized in Table 3 for the 208 eyes that attended Month 24. At least one ocular AE occurred in 32 Cohort 1 eyes (18.2%) and 9 Cohort 2 eyes (28.1%). Rates of procedurally related events were low and comparable between the cohorts. No eye developed clinical hypotony or choroidal effusion or folds. One Cohort 1 eye had anterior rotation of the ciliary body with anterior displacement of the intraocular lens, assessed as probably related to the allograft and managed with atropine. There was one event each of corneal abrasion and retained cortical fragment requiring removal. Loss of ≥ 2 lines of acuity at ≥ 90 days occurred in 8 eyes (3.8%) and confirmed visual field worsening ≥ 2.5 dB in 9 eyes (4.3%). BCVA improved from 0.38 ± 0.50 logMAR at baseline to 0.08 ± 0.18 at Month 24 in Cohort 1 (N = 127; mean change: −0.30 ± 0.48; p < 0.001) and from 0.43 ± 0.66 logMAR at baseline to 0.27 ± 0.28 at Month 24 in Cohort 2 (N = 23; mean change: −0.16 ± 0.64; p = 0.23).

**Table 3.**
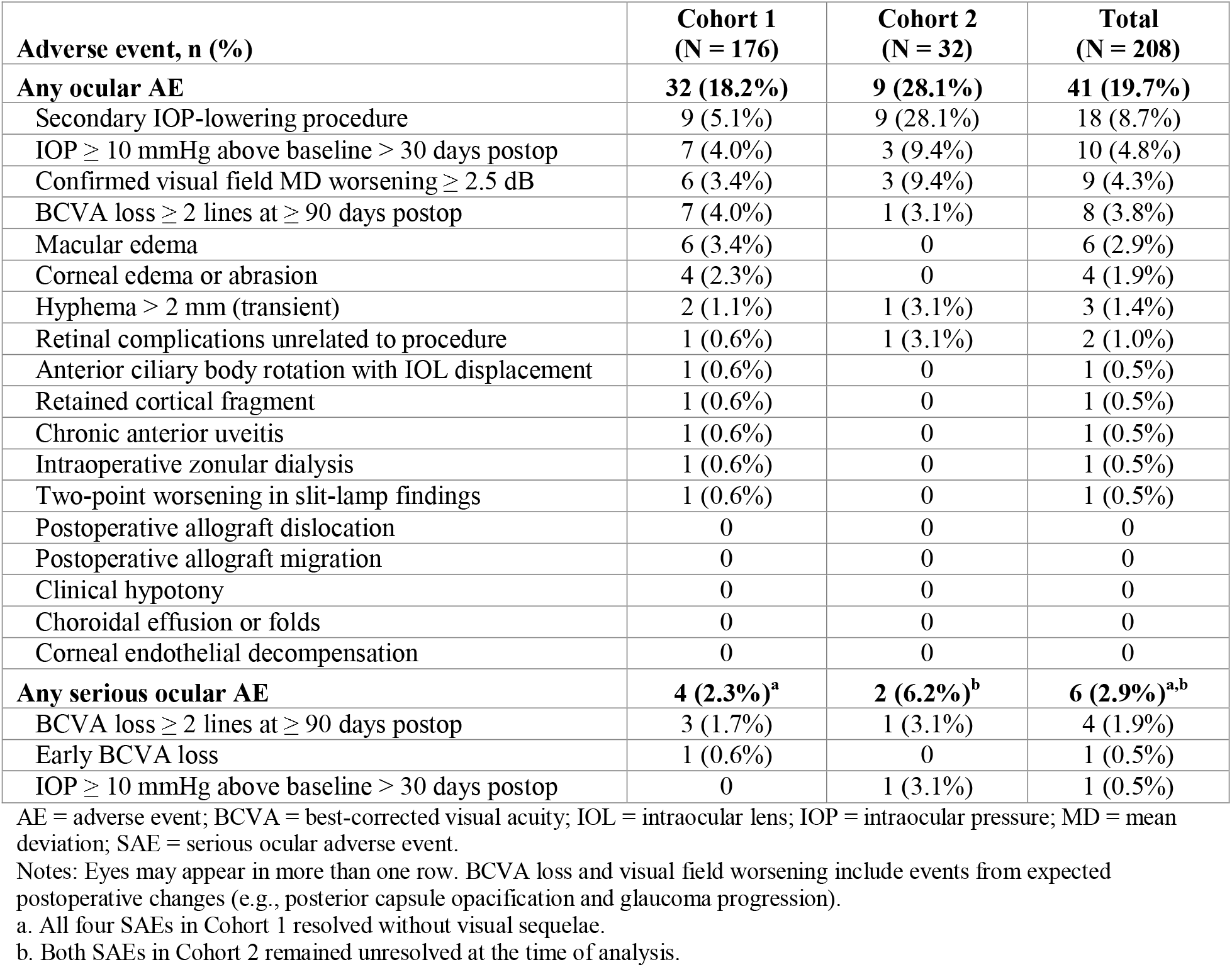
Ocular adverse events through Month 24.

Serious ocular AEs occurred in 6 eyes: 4 (2.3%) in Cohort 1 and 2 (6.2%) in Cohort 2. These comprised 4 eyes with best-corrected visual acuity (BCVA) loss of ≥ 2 lines measured ≥ 90 days postoperatively, 1 with early BCVA loss, and 1 with IOP elevated ≥ 10 mmHg above baseline. The four serious events in Cohort 1 resolved without visual sequelae; the two serious events in Cohort 2 remained unresolved at the time of analysis. In one, BCVA fell from 20/30 to 20/100 in association with an epiretinal membrane and was judged unrelated to the allograft. In the other, an eye with a baseline IOP of 35 mmHg recorded 45 mmHg on the day after a second cyclophotocoagulation, then settled to 7 mmHg on two medications by Month 24 with the event remaining open on the case report form.

Seriousness was assigned by the investigator using standard regulatory criteria and not by the magnitude or timing of the acuity change. Therefore, the 4 serious BCVA losses appear twice in Table 3: once in the “any ocular AE” category (8 eyes [3.8%]) and again the “any serious ocular AE” category (4 eyes [1.9%]). Likewise, the 1 serious IOP elevation appears twice in Table 3.

There were no reports of secondary allograft migration throughout the analysis period. At Month 24, gonioscopy was performed in 156 eyes. Of these, the allograft was optimally positioned in 140 (89.7%), the cleft was open around the allograft in 18 (11.5%), and no eye had the allograft touching the cornea at any visit, including Month 24. Partial peripheral anterior synechiae (PAS) were recorded in 9 eyes (5.8%) and total synechiae in 6 (3.8%).

In the limited subset of evaluable patients with real-world specular microscopy, an exploratory analysis revealed a rate of ECD loss below the 10% threshold anticipated for phaco-MIGS [16, 17]: 8.4% by Month 24 in the 34 Cohort 1 eyes (from a baseline of 2,415 ± 385 to 2,219 ± 430 cells/mm²) and 3.0% in the 3 Cohort 2 eyes (from a baseline of 2,547 ± 373 to 2,458 ± 338 cells/mm²). Corneal edema occurred in 3 eyes, all resolved, and no eye developed corneal decompensation.

## Discussion

### Efficacy

In this multicenter registry, allograft-reinforced cyclodialysis lowered both IOP and medication burden on average in both cohorts. The IOP-lowering effect was more pronounced in the standalone population (Cohort 2), where baseline pressure was much higher than in the phaco-combined cohort (Cohort 1), in which eyes were mostly controlled and the therapeutic objective is often medication reduction.

When combined with phacoemulsification, the procedure lowered medicated IOP by 3.1 mmHg and medication use by 0.5 from a mean baseline of 18.0 mmHg; performed standalone, it lowered IOP by 6.4 mmHg (27.1%) and medication use by 1.2 from 20.0 mmHg in eyes managed without further surgery. Nearly half of phaco-combined eyes and three in ten standalone eyes were controlled on no glaucoma medication at 24 months. No eye developed clinically significant hypotony or choroidal effusion. More eyes in the standalone cohort needed secondary incisional surgery for optimal IOP control.

The two cohorts represent distinct, clinically selected real-world populations. At baseline, the standalone population (Cohort 2) had higher mean medicated IOP than the phaco-combined population (Cohort 1; 20.8 versus 17.9 mmHg) despite approximately twice as many glaucoma medications (2.9 versus 1.4 agents); 56.3% versus 23.9% of eyes had baseline IOP ≥21 mmHg. These differences fit a paradigm in which intervention during cataract surgery often reduces medication burden while maintaining pressure control, whereas standalone intervention addresses the need for additional IOP lowering despite substantial medical therapy. Similar selection patterns are reported for other MIGS procedures: in the iStent inject series of Guedes et al., standalone eyes had higher baseline IOP and medication use (17.8 mmHg on 2.60 medications versus 16.0 mmHg on 2.14) [18], and Hengerer et al. reported greater baseline pressure, medication burden, and prior glaucoma procedures in standalone eyes [19]. Sieck et al. found the same pattern with Kahook Dual Blade goniotomy: 20.4 mmHg on 3.1 medications in standalone eyes versus 16.7 mmHg on 1.9 in combined eyes [20]. Outcomes also depend on baseline pressure: in the phaco-Hydrus study of Ang et al., normal-tension eyes starting at 13.5 mmHg sustained medication reduction without significant further IOP lowering [21]. In the phaco-combined OMNI canaloplasty–trabeculotomy arm of ROMEO, eyes with baseline IOP ≤18 mmHg maintained pressure control (14.1 to 13.4 mmHg) while medications fell from 1.6 to 0.9, whereas eyes above 18 mmHg reduced both IOP (21.9 to 15.1 mmHg) and medications (2.0 to 1.1) at 12 months [22]. Collectively, these findings support interpreting the CREST cohorts according to their differing baseline treatment needs, rather than attributing their differing outcomes to surgical setting alone.

Safety was comparable to canal-based interventions (e.g., iStent, canaloplasty, goniotomy) for postoperative hyphema, IOP spikes, and other key AEs, including the low rates of serious AEs, which, as anticipated, were more frequent in the more severe standalone group (Cohort 2) than the mild-to-moderate phaco-combined group (Cohort 1).

Table 4 places the CREST findings alongside published 24-month MIGS outcomes; medicated real-world studies are the most relevant comparators. CREST measured medicated IOP, whereas pivotal randomized trials used medication washout and higher untreated baseline pressures. In the iStent inject and HORIZON trials, phacoemulsification alone reduced unmedicated diurnal IOP by 5.4 and 5.3 mmHg, respectively, with incremental device effects of 1.6 and 2.3 mmHg [6, 7]. Medication status materially affects observed outcomes: a meta-analysis of phacoemulsification-alone trial arms reported 12-month reductions of 5.25 mmHg with washout versus 3.13 mmHg without, falling to 1.81 mmHg after excluding an outlier [3]. Because medication status, baseline pressure, and patient selection influence measured reductions, subtracting a cataract-surgery effect from another population would not isolate AlloFlo’s contribution. Its incremental efficacy in combined surgery therefore remains undetermined.

**Table 4.** Published 24-month IOP outcomes alongside present study results.

| **Study Description** | **N** | **IOP state** | **Δ IOP** | **Follow-up** | **Efficacy Notes** | **Safety Notes** | **Source** |
|---|---|---|---|---|---|---|---|
| **Phacoemulsification alone** |  |  |  |  |  |  |  |
| Retrospective RWE study of EHRs | 12,240 | Medicated | −1.2 (DiD) | 1–18M | Matched-cohort study; DiD −2.5 if BL IOP ≥ 20 mmHg | Glaucoma surgery OR 1.48 in POAG | [5] |
| Meta-analysis, OAG | 2,315 | Mixed | −3.8 | 24M | Pooled estimate | Not reported | [4] |
| iStent inject pivotal, control arm | 118 | Washout | −5.4 | 24M | 61.9% ≥ 20%; 50.0% ≤ 18 | Comparable to stent arm | [6] |
| HORIZON, control arm | 187 | Washout | −5.3 | 24M | Eligibility: IOP 22–34 mmHg | No significant difference vs stent arm | [7] |
| COMPASS, control arm | 131 | Washout | −5.4 | 24M | 60% ≥ 20% | > 98% BCVA ≥ 20/40 | [32] |
| **Phacoemulsification + trabecular or canal-based MIGS** |  |  |  |  |  |  |  |
| iStent inject, FGB registry | 224 | Medicated | −3.1 | 24M | Matched analysis | 5.4% secondary procedure | [24] |
| Hydrus, FGB registry | 120 | Medicated | −2.3 | 24M | Same study as above | 7.5% secondary procedure | [24] |
| Kahook Dual Blade | 51 | Medicated | −8.4 | 24M | Medications 3.3 → 2.3 | 12% secondary procedure | [33] |
| iStent inject pivotal | 387 | Washout | −7.0 | 24M | 75.8% ≥ 20%; 63.2% ≤ 18 | Comparable to control | [6] |
| Hydrus, HORIZON | 369 | Washout | −7.6 | 24M | 77.3% ≥ 20% | No serious device-related AEs | [7] |
| **Phacoemulsification + supraciliary procedure** |  |  |  |  |  |  |  |
| CyPass, COMPASS | 374 | Washout | −7.4 | 24M | 77% ≥ 20% | Withdrawn for endothelial cell loss | [32, 34] |
| MINIject | 27 | Medicated | −3.8 | 24M | Medications 2.11 → 0.38 | 16.2% AEs; no further procedures | [35] |
| Allograft-reinforced cyclodialysis with AlloFlo, single-center | 31 | Medicated | −8.1 | 24M | BL IOP 21.9 mmHg on 1.2 medications | No serious ocular AEs | [11] |
| Present study, Cohort 1 | 167 | Medicated | −3.1 | 24M | 44.9% ≥ 20%; 86.2% ≤ 18 | 5.1% secondary procedure; 1.1% reoperation (KM) | Present study |
| Present study, BL IOP ≥ 21 mmHg | 40 | Medicated | −9.2 | 24M | 36.1% IOP reduction with no change in meds | Not reported separately from full Cohort 1 | Present study |
| **Standalone glaucoma surgery** |  |  |  |  |  |  |  |
| iStent infinite | 72 | Medicated | −5.9 | 12M | Refractory OAG; 12-mo endpoint | No explant, infection, device-related interventions or hypotony | [36] |
| OMNI | 38 | Washout | −10.0 | 24M | Not comparable with medicated data | 44.7% hyphema (all resolved); 5.3% secondary procedure | [37] |
| XEN 45, FGB registry | 646 | Medicated | −4.6 | 24M | Pooled standalone and combined | 18% secondary procedure | [38] |
| PRESERFLO MicroShunt | 81 | Medicated | −7.6 | 24M | Bleb-forming, with MMC | 7.4% reoperation; 25.9% IOP rise needing treatment | [39] |
| Present study, Cohort 2 | 23 | Medicated | −6.4 | 24M | −27.1% IOP and −1.2 medications from BL | 28.1% secondary procedure; 15.6% reoperation (KM) | Present study |

AE = adverse event; BCVA = best-corrected visual acuity; BL = baseline; DiD = difference in differences; EHR = electronic health record; FGB = Fight Glaucoma Blindness!; IOP = intraocular pressure; KM = Kaplan–Meier; MIGS = minimally invasive glaucoma surgery; MMC = mitomycin C; OAG = open-angle glaucoma; OR = odds ratio; POAG = primary open-angle glaucoma; RWE = real-world evidence.
Notes: Washout studies report unmedicated diurnal IOP after medication washout; medicated studies report IOP on the eye’s current regimen. The two are not interchangeable and washout designs yield larger changes in both arms. Safety entries are as reported by each source; definitions of reoperation, secondary procedure and complications differ between studies, and rates track baseline disease severity. They are shown for context and are not directly comparable.

Within medicated real-world data, Cohort 1’s 3.1-mmHg reduction from 18.0 mmHg, with a 0.5-agent medication reduction, is numerically consistent with established phaco-MIGS outcomes. In a 340-eye multicenter phaco-iStent inject series, IOP decreased from 16.4 to 13.7 mmHg and medications from 1.49 to 0.49 at 24 months [23]. Propensity-matched Fight Glaucoma Blindness registry cohorts showed reductions of 3.1 mmHg with iStent inject and 2.3 mmHg with Hydrus, with medication reductions of 1.0 and 0.5 agents [24]. Larger reductions are reported in canaloplasty populations starting higher: Gallardo’s mixed standalone and phaco-combined iTrack series showed IOP falling from 20.0 to 13.5 mmHg and medications from 2.77 to 1.7 [25]. In the two-year ROMEO extension, OMNI canaloplasty–trabeculotomy reduced IOP by 6.1 mmHg in combined eyes and 7.4 mmHg in standalone eyes starting above 18 mmHg, compared with 0.6 and 2.3 mmHg in the respective lower-pressure groups; medications decreased across all four subgroups [26]. These observations support interpreting pressure lowering jointly with baseline IOP and medication reduction, while recognizing that baseline-stratified changes may also reflect regression to the mean.

Cohort 2 addressed a different treatment context, with higher baseline medicated IOP despite greater medication use. Its primary 24-month reduction of 6.4 mmHg from 20.0 mmHg, with a 1.2-agent medication reduction, falls within the numerical range reported for real-world standalone canal-based surgery. IOP and medication measurements recorded after secondary IOP-lowering procedures were excluded from efficacy analyses, so subsequent interventions did not contribute to the reported reductions. Secondary procedures occurred in 28.1% of standalone versus 5.1% of combined eyes, while Kaplan–Meier estimates of incisional reoperation were 15.6% and 1.1%, respectively. A similar direction of association was observed in the IRIS Registry, where two-year reoperation estimates were 24% after standalone goniotomy/canaloplasty versus 6% with concurrent phacoemulsification [27]. Differences in patient selection, ascertainment, and reoperation definitions preclude direct comparisons of these rates.

Standalone eyes requiring further intervention had higher baseline IOP than those remaining in the primary efficacy analysis (22.8 versus 20.0 mmHg). The primary estimate consequently describes outcomes while eyes remained managed with the index procedure and medications, rather than outcomes across the entire original cohort. In the failure-inclusive sensitivity analysis, these eyes were assigned their baseline IOP and medication counts, yielding a more conservative standalone reduction of 4.6 mmHg, with 50% achieving ≥20% reduction. The findings therefore support sustained IOP and medication reductions through 24 months in eyes managed without further procedures, alongside a clinically relevant need for additional intervention in about one-quarter of standalone eyes. Comparisons with other MIGS provide context but do not establish superiority or equivalence, and the combined-surgery findings cannot distinguish the contribution of allograft-reinforced cyclodialysis from that of phacoemulsification.

### Safety

The observed safety profile of allograft-reinforced cyclodialysis was broadly consistent with established canal-based MIGS, for which hyphema, transient IOP elevation, and anterior-segment inflammation are characteristic postoperative events. A meta-analysis of 95 studies encompassing 9,733 eyes found no increase in sight-threatening events with phaco-combined MIGS compared with phacoemulsification alone, although pooled hyphema rates varied by mechanism: 3.5% for trabecular bypass, 15.5% for non-GATT goniotomy, and 27.7% for GATT [28]. Phaco-combined OMNI canaloplasty–trabeculotomy produced hyphema, IOP spikes, and mild inflammation in 4%, 5%, and 11% of eyes [22], compared with 4%, 6%, and 13% after standalone OMNI surgery [29]. Kahook Dual Blade goniotomy was associated with transient Day-1 hyphema in 17.3% and Week-1 IOP elevations >10 mmHg in 10.2% [20], whereas the iStent inject pivotal trial demonstrated overall safety similar to cataract surgery alone through 24 months [6].

In the present CREST analysis, no clinically significant hypotony, choroidal effusion or folds, or corneal decompensation occurred, and no allograft–cornea contact was recorded at Month 24 gonioscopy. Prior allograft-reinforced cyclodialysis reports provide supportive context: among 243 eyes, postoperative hyphema occurred in 2% and resolved conservatively; one prolonged iritis event and one numeric hypotony event without maculopathy also resolved [10]. A separate 12-month standalone CREST analysis reported no persistent inflammation, allograft rejection, clinically significant hyphema, or scaffold migration, although IOP elevations ≥10 mmHg occurred in 17%, most resolving within 30 days [12]. These earlier findings should not be extrapolated to imply absence of such events throughout the present 24-month cohort.

Overall ocular adverse events, serious ocular adverse events, and protocol-defined IOP elevations were numerically more frequent in standalone than phaco-combined CREST eyes (28.1% versus 18.2%, 6.2% versus 2.3%, and 9.4% versus 4.0%, respectively). Six eyes experienced serious ocular events, including two standalone events unresolved at the analysis cutoff. Although procedure-related event rates were low and similar between cohorts, the contribution of baseline disease burden to these differences cannot be determined from this unmatched observational analysis. The findings therefore support a favorable observed safety profile, without establishing equivalence to other MIGS. In a review of 401 studies encompassing 39,381 eyes, inconsistent adverse-event definitions and incomplete capture of late endothelial loss and peripheral anterior synechiae materially limited cross-procedure comparisons; standardized reporting and longer prospective follow-up remain necessary [30]. Protocol-defined IOP elevations clustered early, and the mechanisms most likely to account for them are retained ophthalmic viscosurgical device, postoperative inflammation with fibrin occluding the cleft, and corticosteroid response, rather than the allograft itself.

At Month 24, partial and total synechiae were recorded in 9 (5.8%) and 6 (3.8%) of 156 evaluable eyes, and no allograft–cornea contact was recorded at any time. For comparison, PAS occurred in 14.6% of Hydrus eyes during five-year HORIZON follow-up, without serious sequelae attributable to the Hydrus-associated adhesions [31]. These comparisons remain descriptive because cyclodialysis-cleft adhesions may differ anatomically and functionally from implant-associated PAS, and ascertainment and follow-up were not harmonized. In the CREST specular-microscopy subset, endothelial cell density declined by 8.4% at 24 months in 34 phaco-combined eyes, with no detectable change in standalone eyes. This loss was numerically lower than the 13.1% with phaco-iStent inject and 12.3% with phacoemulsification alone in the pivotal trial, although the small, real-world CREST subset, without a reading center or protocolized specular microscopy, precludes inference of superior endothelial preservation [6]. The absence of corneal decompensation and of recorded allograft–cornea contact may partly reflect the compliant, homologous acellular scleral tissue and its placement within the supraciliary cleft, clear of the corneal endothelium. Compared with exogenous hardware, this tissue-based construct offers a plausible rationale for reduced mechanical irritation and foreign-body response; however, neither mechanism was directly measured, and material composition cannot be separated from positioning, geometry, or surgical technique. The results therefore support the biological rationale for allograft reinforcement without establishing superior biocompatibility or corneal and angle safety. Optional endothelial measurements, small evaluable subsets, and nonstandardized synechiae characterization warrant longer follow-up with serial specular microscopy and anatomically defined gonioscopic grading. The cleft was recorded as open around the allograft in 18 eyes (11.5%) at Month 24. Because no eye had allograft-cornea contact and the allograft was optimally positioned in 89.7%, this appearance is more consistent with a cleft that remains patent beyond the allograft footprint, and therefore with continued uveoscleral flow, than with displacement of the tissue. The converse finding of partial or total synechiae in 15 eyes may represent gradual remodeling of the cleft margins. Both readings are descriptive because gonioscopic appearance was not graded against a predefined anatomic scale.

No secondary allograft migration was reported through 24 months. This may reflect the compliant scleral tissue conforming to the endoscleral wall, with close tissue apposition and frictional engagement as plausible mechanisms of retention, although surface friction and fixation strength were not measured, and greater resistance to migration than metal or polymer implants has not been established.

### Strengths and Limitations

This is the largest multicenter report of 24-month safety and effectiveness for allograft-reinforced cyclodialysis, and it draws on prospectively collected data with protocol-defined adverse event categories.

Several limitations qualify these findings. The design is observational and single-arm, so in the combined cohort the contribution of the allograft procedure cannot be separated from that of phacoemulsification; only a randomized comparison can do that. IOP was measured on medication without washout, so these values are not comparable with pivotal trials that used washout. The two cohorts differed substantially in baseline pressure, medication burden and disease severity, were not matched, and are therefore reported separately without a formal between-cohort test of effectiveness. Stratification by baseline pressure was exploratory and unadjusted for multiplicity, and because tier membership is defined by the same measurement from which change is calculated, tier-level changes remain susceptible to regression to the mean. The standalone cohort comprised 23 eyes in the primary analysis, which limits the precision of every estimate derived from it, and the 9 eyes that underwent a secondary procedure had higher baseline pressure, so the failure-inclusive estimate is the more conservative description. Specular microscopy was optional, as real-world standard of care does not include ECD biometry for glaucoma procedures, so endothelial estimates rest on a limited subset of eyes. Lastly, both eyes of 50 patients were included and analyzed at the eye level without adjustment for inter-eye correlation (baseline IOP r = 0.73, Month 24 r = 0.56), so confidence intervals may be modestly narrow. Follow-up beyond 24 months is not yet available, and the durability of cleft patency over longer intervals remains unknown.

## Conclusions

Allograft-reinforced cyclodialysis, performed alone or with phacoemulsification, was associated with significant reductions in medicated IOP and medication burden sustained through 2 years, with an increased proportion of eyes requiring no medication and a favorable observed safety profile consistent with a minimally invasive ab-interno intervention.

## Acknowledgments

We thank the participants of the study.

## Medical Writing and Editorial Assistance

No medical writing or editorial assistance was received in the preparation of this article.

## Data Availability

The datasets generated during and/or analyzed during the current study are available from the corresponding author on reasonable request, subject to a data use agreement.

## Authorship

All named authors meet the International Committee of Medical Journal Editors (ICMJE) criteria for authorship of this article, take responsibility for the integrity of the work as a whole, and have given their approval for this version to be published.

## Ethical Approval

This study was performed in accordance with the ethical standards of the 1964 Declaration of Helsinki and its later amendments. The CREST protocol was approved by Advarra Institutional Review Board (approval MOD02608603), and all participants provided written informed consent.

## Author Contribution

Conceptualization: Tsontcho Ianchulev and Robert N. Weinreb. Methodology and formal analysis: Tsontcho Ianchulev. Investigation and data acquisition: all authors. Writing, original draft preparation: Robert N. Weinreb and Tsontcho Ianchulev. Writing, review and editing: all authors. All authors read and approved the final manuscript.

